# Validation of an MRI-based classification of peroneus brevis tendon morphology: a four-type system with high inter-rater reliability for sports imaging

**DOI:** 10.1101/2025.05.18.25327845

**Authors:** Rafał Zych, Dan Mocanu, Ymer Hagberg, Katarzyna Bokwa-Dąbrowska, Michael Huuskonen, Isaac Romanus, Dawid Dziedzic, Pawel Szaro

**Affiliations:** Department of Clinical and Descriptive Anatomy, Medical University of Warsaw, Poland; Department of Musculoskeletal Radiology, Sahlgrenska University Hospital, Gothenburg, Sweden; Department of Radiology, Institute of Clinical Sciences, Sahlgrenska Academy, University of Gothenburg, Gothenburg, Sweden

**Author notes:** **Corresponding author** Pawel Szaro, Address: Radiology, Göteborgsvägen 31, 431 80 Göteborg, Sweden.

**Keywords:** MRI-based classification, peroneus brevis tendon, tendon morphology, sports imaging, inter-rater agreement, magnetic resonance imaging

## Abstract

**Purpose:** Split tears of the peroneus brevis tendon are a common and often underrecognized cause of lateral ankle pain and instability in athletes. Although MRI is widely used for ankle assessment, no validated system exists for classifying normal morphological variation of the peroneus brevis tendon. This study aimed to validate an MRI-based classification system.

**Methods:** We analysed 130 normal peroneus brevis tendons (power > 0.8, α = 0.05, effect size = 0.31) on consecutive 3T ankle MRI scans. Persons with fractures, ligament injuries, or peroneal tendon pathology were excluded. Seven independent raters classified tendons into four types: general flat, flat with lateral bulge, flat with medial bulge, and oval. Inter-rater reliability was assessed using Cohen’s kappa, Gwet’s AC1, and Fleiss’ kappa. Classification robustness was evaluated using F1 scores, ROC analysis, and precision-recall metrics.

**Results:** The most common forms were general flat (37.7%) and oval (22.3%). Fleiss’ kappa was 0.691 (95% CI 0.629–0.752), with strong pairwise agreement (Cohen’s kappa and Gwet’s AC1: 0.58–0.87). ROC AUC was 0.89; majority voting improved agreement (F1 = 0.93).

**Conclusion:** This classification system is robust and reliable, supporting consistent MRI-based assessment of tendon morphology in clinical and research settings.

## INTRODUCTION

Peroneus brevis split tears are a common but frequently underdiagnosed cause of chronic lateral ankle pain and instability in athletes[1-5]. These injuries often arise after ankle sprains and may persist for years if not properly recognized, compromising performance and delaying return to sport[6]. Variations in tendon morphology, particularly flattening, may predispose to split tears by increasing mechanical stress within the retromalleolar groove[7]. Despite this, no validated classification of peroneus brevis tendon shape exists at the level where split tears typically originate[8]. A reliable, reproducible classification system is therefore essential to improve diagnostic accuracy, guide treatment, and support early return to sport. Previous studies have used terms such as “chevron” or “bifid” [8, 9] to describe the cross-sectional shape on transverse magnetic resonance imaging (MRI), suggesting that an abnormal tendon shape may be an important imaging biomarker for split tear, which can be difficult to detect on MRI.

Despite the clinical significance, peroneus brevis split tear remains challenging due to the complex anatomical relationship of the peroneal tendons and the inherent limitations of the available imaging modalities [10]. The peroneus brevis and peroneus longus tendons lie adjacent to each other, tightly confined within the retromalleolar groove and stabilised posteriorly by the superior peroneal retinaculum [11, 12]. Tight anatomical relationships between the content of the superior peroneal tunnel [13] make it difficult to assess peroneus brevis pathology on ultrasound and MRI.

Peroneus brevis split tears are much more common than lacerations[14] and may vary, ranging from complete tears, where the tendon is fully divided into two parts, to non-complete tears. Non-complete tears are particularly difficult to detect with imaging, as their radiological features may overlap with normal anatomical variations. One proposed imaging marker is an alteration in tendon shape on transverse cross section on MRI[8]. Despite clinical observations, there is no consensus in the literature regarding the normal anatomical variability of the peroneus brevis tendon. Its shape has been hypothesised to influence the susceptibility to split tears, with some authors suggesting that flatter peroneus brevis tendons are at greater risk due to compression by the peroneus longus against the retromalleolar sulcus[15]. However, there is no validated classification system for peroneus brevis morphology, nor any systematic studies evaluating whether anatomical variants contribute to the risk of split tear formation.

In contrast, the morphology of the retromalleolar groove, a low-lying muscle belly, or the presence of the peroneus quartus has been studied extensively [12, 16-20]. A shallow sulcus and sharp contour have been linked to peroneal tendon instability, emphasizing the role of anatomical predisposition in tendon pathology. While other anatomical variations have been studied, little attention has been given to the anatomical variability of the peroneus brevis tendon.

The lack of a standardised classification system for the peroneus brevis tendon morphology represents a significant gap in musculoskeletal radiology. Given the importance of reliable and reproducible imaging criteria, any proposed classification must undergo rigorous validation to ensure its applicability in both clinical and research settings. To our knowledge, no studies have systematically classified peroneus brevis tendon morphology on MRI. Thus, we addressed this gap by proposing a classification system of peroneus brevis tendon shape variability and evaluating its potential clinical implications. We aimed to assess the reliability of our proposed peroneus brevis tendon classification system, to identify potential misclassification patterns, and to explore factors that may influence classification accuracy.

## METHODS

### Study design and observer validation framework

This study evaluated the inter-rater agreement regarding a proposed classification of the peroneus brevis tendon form on the transverse section at the level of the lateral malleolus. This study was conducted and reported in accordance with the Guidelines for Reporting Reliability and Agreement Studies (GRRAS) [21].

### MRI examinations

The dataset was compiled retrospectively from MRI examinations performed our institution in 2021–2024 with a 3T machine due to sports injuries or pain related to physical activity. We retrospectively reviewed 439 ankle MRI examinations performed at our institution between January 2021 and December 2024. All examinations followed our standard ankle MRI protocol and had been conducted due to trauma or pain in the ankle. Only one MRI per patient was included. Exclusion criteria encompassed peroneal tendon pathology, fractures, postoperative changes, image artifacts, and incomplete clinical data (**Supplementary material Figure S1**).

### Raters

A structured visual assessment approach was used for grading. Seven independent raters with different backgrounds (2 musculoskeletal radiologists with 7 and 10 years of experience in musculoskeletal radiology, 1 radiology resident, 1 medical doctor, 2 physiotherapists, and 1 fifth-year medicine student), visually assessed each tendon and assigned one of four predefined variations based on morphology. The classification system was developed from a preliminary study by our team and refined through pilot consensus rounds. To establish a reference standard, a consensus classification agreement among musculoskeletal radiologists. To assess inter-rater reliability, each rater independently classified all cases in a blinded manner. Intra-rater reliability was assessed after a washout period. Because assessing intra-rater variability using a subset of data is a valid approach that has been used in previous studies [22], a subset comprising 30% of the subjects was randomly selected by an individual not involved in the study. The same raters re-evaluated these cases after a 3-week washout period.

### Data acquisition and preprocessing

#### Sampling

Convenience sampling was used to select all available MRI scans of the ankle that met the inclusion criteria. A power analysis was performed prior to the study to determine the sample size. With power > 0.8, alpha = 0.05, an effect size of 0.31 based on our preliminary study and a 10% buffer, 130 participants were required.

#### Preprocessing

Analysis of the peroneus brevis tendon shape was based on proton density–weighted axial images. A senior musculoskeletal radiologist selected a representative image at the level of the lateral malleolus, inferior to the syndesmosis, at the level of the ankle joint space. This level was chosen based on our preliminary study, which demonstrated that the tendon shape remains consistent within this region. The selected images were exported, and an electronic form was created, including the images and four classification options (general flat, flat with a medial bulge, flat with a lateral bulge, and oval tendon), which was then distributed to the raters. Each rater received detailed instructions including definitions of tendon forms and examples (not including the evaluated cases) regarding tendon forms and assessed tendons using a standardised protocol for the study.

### Proposed classification

The proposed classification (nominal variable) is based on the visual assessment of the ratio of thickness (the shortest diameter) to the width of the tendon (the longest diameter). The four types are defined below (Fig. 1).

**Fig. 1.**
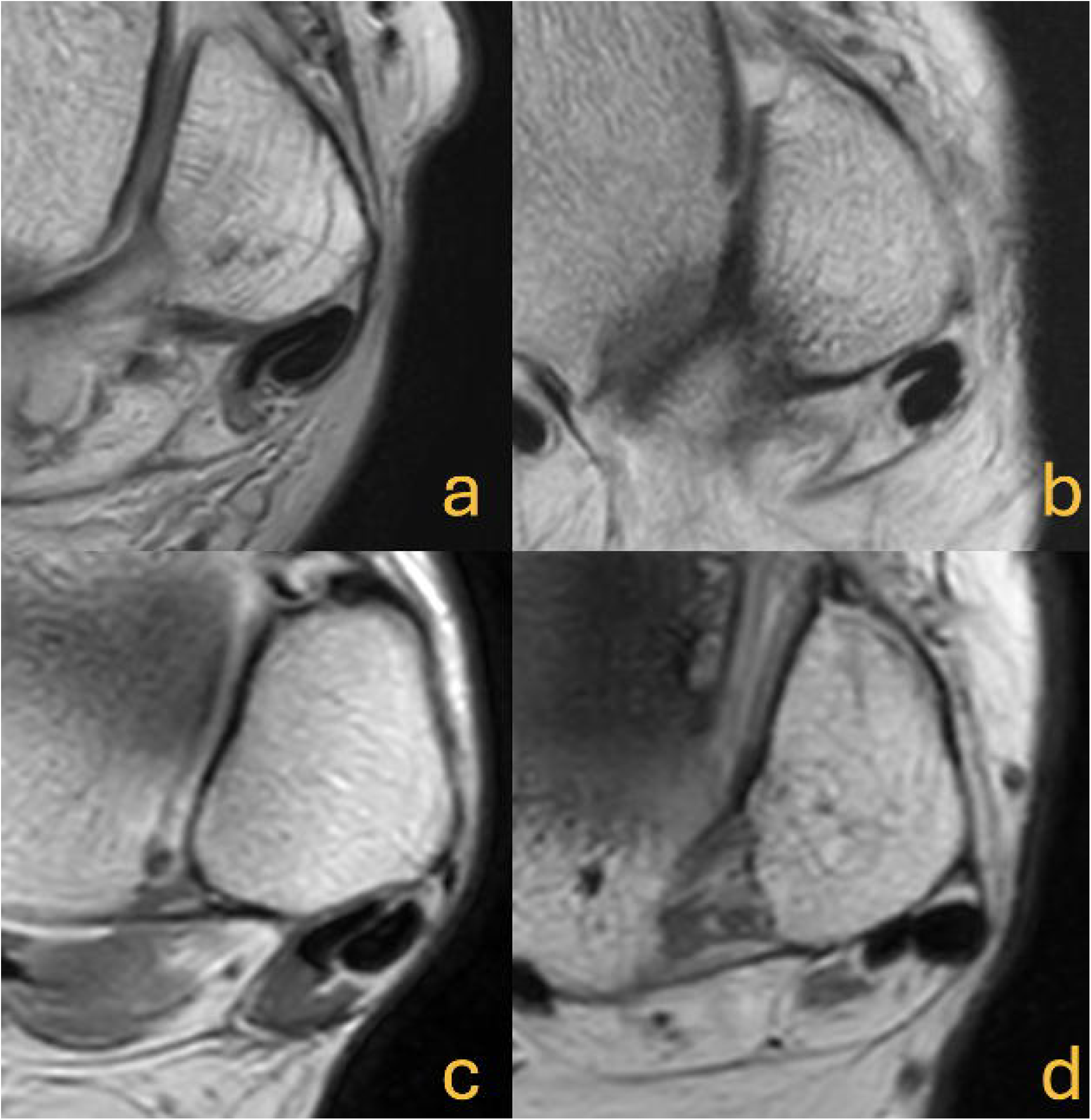
Axial section at the level of the lateral malleolus and the ankle joint space (magnetic resonance images with proton-density weighting, left sides). The four morphological types of the peroneus brevis tendon are: **A** general flat, **B** flat with a lateral bulge, **C** flat with a medial bulge, and **D** oval tendon

#### General flat tendon

The thickness of the tendon (defined as the anteroposterior [AP] dimension) is relatively consistent across its length. The width of the tendon is significantly greater than its thickness. The tendon in this form can be either straight, curved, or folded.

#### General flat with a lateral bulge

The tendon is flattened, but the lateral edge bulges, increasing the thickness in the lateral part.

#### General flat with a medial bulge

The tendon is flattened, but the medial edge bulges, resulting in an increase in thickness in the medial part.

#### Oval tendon

The peroneus brevis tendon takes the shape of an oval, resembling the cross-section of the peroneus longus tendon. The thickness is slightly smaller than the width.

### Gold standard

The final result was a consensus between two musculoskeletal radiologists, which was considered to be the gold standard.

### Statistical analysis

R version 4.4.3 (R Core Team, Vienna, Austria) and RStudio version 2024.12.1+563 (2024.12.1+563) were used for statistical analysis and data visualisation. The ggplot2 package was used to visualise the data. Inter-rater agreement across the seven raters was assessed based on Fleiss’ kappa with a bootstrapped 95% confidence interval. For pairwise agreement, Cohen’s kappa and Gwet’s AC1 were calculated between all rater pairs. These statistics were also used to assess intra-rater reliability after a 3-week washout period. Gwet’s AC1 was included as a robust alternative to Cohen’s kappa, particularly in light of potential prevalence and marginal distribution imbalances.

The F1 score, defined as the harmonic mean of precision and recall, was calculated for each rater to assess the performance of the proposed classification. This metric reflects the balance between correctly identifying tendon classifications (precision) and detecting all cases of a given class (recall). The F2 score (β = 2) was calculated to account for cases where recall was prioritised. In addition, a majority vote approach was applied, where the most frequently assigned tendon form among the seven raters was selected as the predicted class. Then, the majority vote classifications were compared to the gold standard (defined as a consensus between two musculoskeletal radiologists) to calculate the overall classification performance metrics.

Receiver operating characteristic (ROC) curve analysis was conducted by using a cumulative one-versus-rest strategy, comparing each tendon form against all others. Macro- averaged values for precision, recall, the F1 score, and the area under the curve (ROC-AUC) are reported to ensure balanced evaluation across all tendon categories.

To identify misclassification patterns, confusion matrices were generated for each rater and for the majority vote. These matrices were used to visualise common errors and to highlight areas of disagreement or uncertainty between raters.

### Ethical considerations

All procedures performed in studies involving human participants were in accordance with the ethical standards of the institutional and/or national research committee and with the 1964 Helsinki Declaration and its later amendments or comparable ethical standards. The National Ethical Authority approved this study and waived informed consent (Dnr 2024-07283-02).

## RESULTS

### Study cohort

A total of 439 ankle MRI examinations were reviewed. After applying the exclusion criteria, 310 examinations were excluded. The final study population comprised 130 examinations performed using our standard ankle protocol for evaluation of trauma or ankle pain (**Supplementary material Figure S1**). The study cohort consisted of 130 persons with a mean age of 39.7 years and a standard deviation of 13.4 years (range: 18–64 years). There were 73 males (56.2%) and 57 females (43.8%). The left side was examined in 69 persons (53.1%), while the right was examined in 61 persons (46.9%).

### Consensus-based tendon classification

The distribution of tendon forms based on the consensus between raters was as follows: 37.7% of cases (n = 49) were classified as general flat, 20.8% (n = 27) as flat with a lateral bulge, 19.2% (n=25) as flat with a medial bulge, and 22.3% (n = 29) as oval tendon. Fig. 1 shows an example of each type.

### Inter-rater agreement

#### Pairwise agreement between raters

Cohen’s kappa – which ranged from 0.589 to 0.865 – and Gwet’s AC1 – which ranged from 0.580 to 0.865 – indicated substantial to almost perfect agreement according to Landis and Koch [23] (Table 1 and **Supplementary material Figure S2-S4**).

**Table 1.**
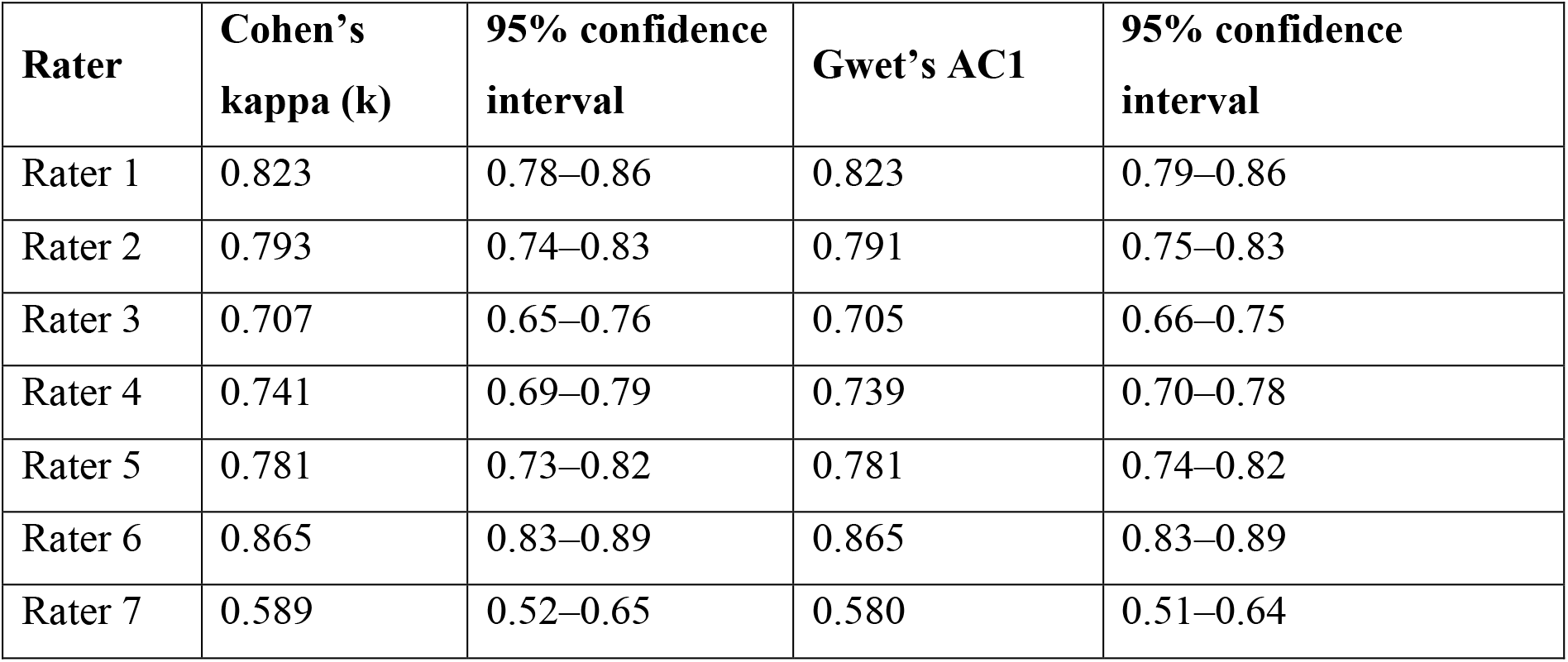
Agreement statistics for each rater compared with the consensus.

| Rater | Cohen's kappa (k) | 95% confidence interval | Gwet's AC1 | 95% confidence interval |
| --- | --- | --- | --- | --- |
| Rater 1 | 0.823 | 0.78–0.86 | 0.823 | 0.79–0.86 |
| Rater 2 | 0.793 | 0.74–0.83 | 0.791 | 0.75–0.83 |
| Rater 3 | 0.707 | 0.65–0.76 | 0.705 | 0.66–0.75 |
| Rater 4 | 0.741 | 0.69–0.79 | 0.739 | 0.70–0.78 |
| Rater 5 | 0.781 | 0.73–0.82 | 0.781 | 0.74–0.82 |
| Rater 6 | 0.865 | 0.83–0.89 | 0.865 | 0.83–0.89 |
| Rater 7 | 0.589 | 0.52–0.65 | 0.580 | 0.51–0.64 |

### Intra-rater reliability

After a 3-week washout period, Cohen’s kappa ranged from 0.57 to 0.93, indicating moderate to almost perfect intra-rater agreement. Gwet’s AC1 was generally higher, ranging from 0.70 to 0.93. Most raters demonstrated substantial to almost perfect reproducibility according to Landis and Koch [23].

### Overall agreement across raters

Fleiss’ kappa across all raters was 0.691 (95% confidence interval 0.629–0.752), indicating substantial agreement according to Landis and Koch [17]. To assess the stability of this result, we performed bootstrapping, resampling the data 1000 times. The 95% confidence interval was 0.629–0.752 (Fig. 2). This narrow confidence interval suggests that the level of agreement between raters is consistent across different resamples. The substantial agreement found, with the lower bound of the confidence interval still above 0.6, indicates reliable rater consistency in classifying the tendon forms.

**Fig. 2.**
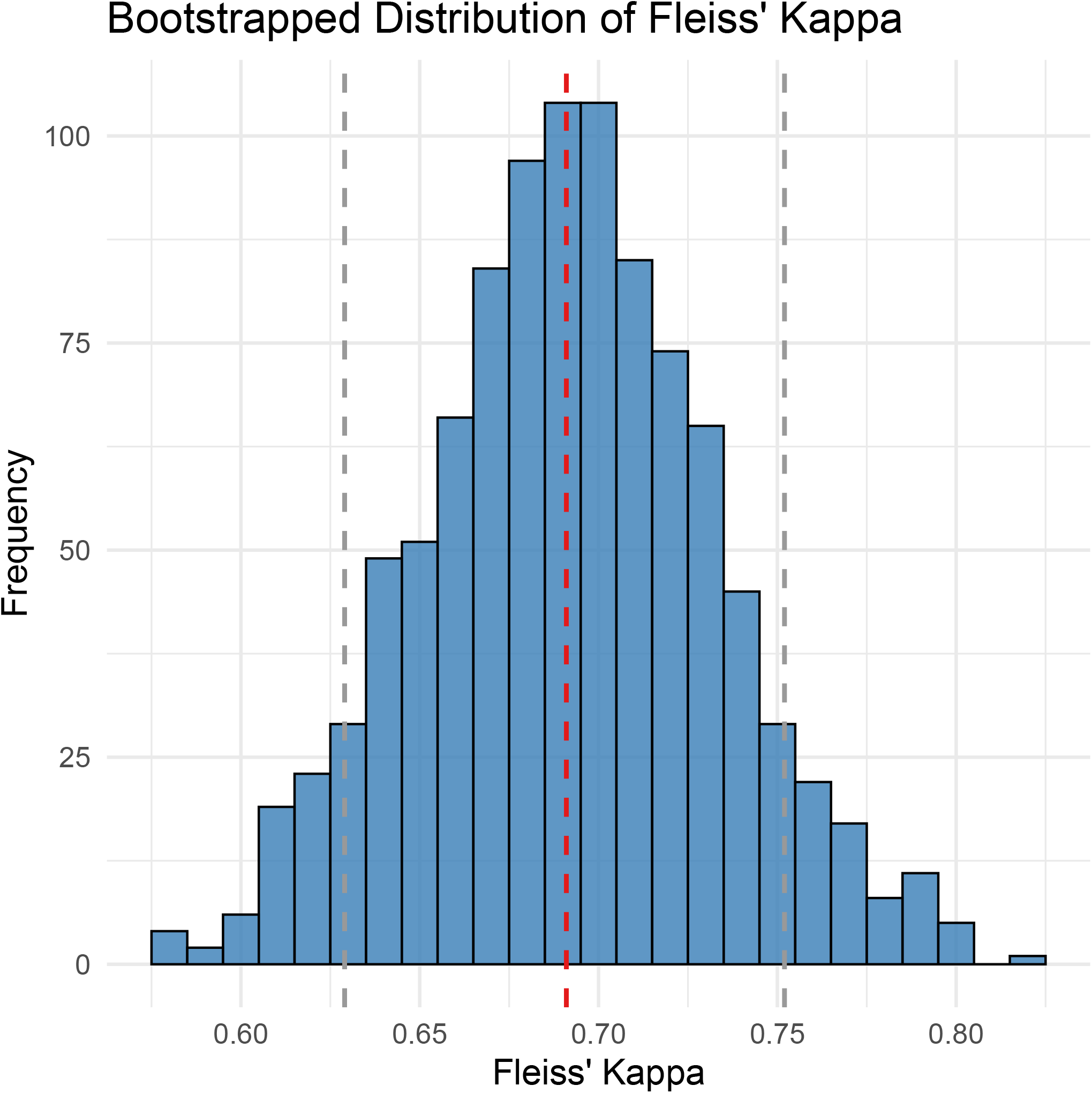
Bootstrapped distribution of Fleiss’ kappa computed from 1,000 resamples. The vertical dashed line represents the mean Fleiss’ kappa (0.691), and the grey dashed lines indicate the 95% confidence interval (0.629–0.752)

### Classification performance metrics

The classification performance of individual raters varied, as reflected in the F1 score, harmonic mean, and F2 score. While most raters demonstrated substantial agreement with the consensus classification, we observed differences in balance precision and recall that affected the overall F1 scores (Table 2). Raters 1, 2, and 5 had consistently high performance, with an F1 score > 0.84, indicating strong agreement with the consensus. Rater 6 exhibited the highest classification accuracy, achieving an F1 score of 0.90, suggesting reliable differentiation between tendon forms. Conversely, Raters 3 and 7 had a lower F1 score (0.78 and 0.68, respectively), indicating greater variability in classification, likely due to challenges in distinguishing between certain tendon forms.

**Table 2.**
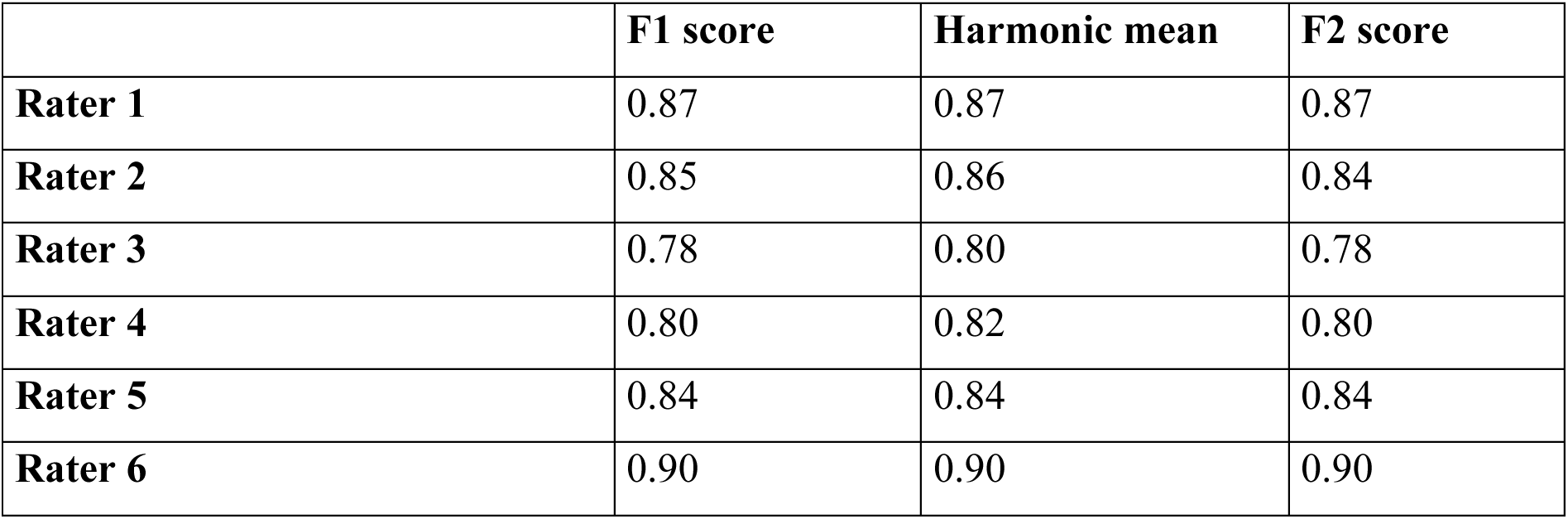
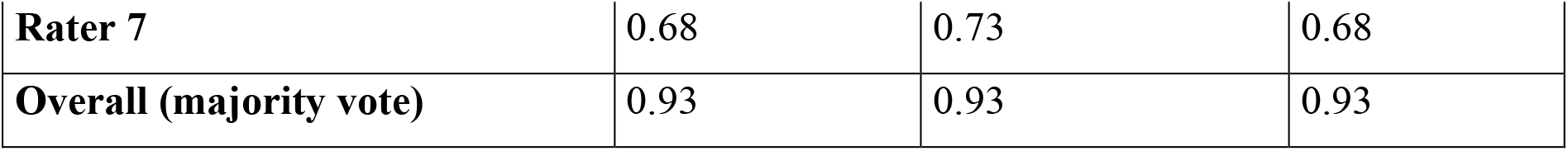
Per-rater and overall performance.

|  | **F1 score** | **Harmonic mean** | **F2 score** |
| --- | --- | --- | --- |
| **Rater 1** | 0.87 | 0.87 | 0.87 |
| **Rater 2** | 0.85 | 0.86 | 0.84 |
| **Rater 3** | 0.78 | 0.80 | 0.78 |
| **Rater 4** | 0.80 | 0.82 | 0.80 |
| **Rater 5** | 0.84 | 0.84 | 0.84 |
| **Rater 6** | 0.90 | 0.90 | 0.90 |
| **Rater 7** | 0.68 | 0.73 | 0.68 |
| **Overall (majority vote)** | 0.93 | 0.93 | 0.93 |

To further assess classification reliability, we applied a majority vote approach, using the most frequently assigned class among all raters as the final prediction (Table 2). This method significantly improved classification performance, achieving an F1 score of 0.93, a harmonic mean of 0.935, and an F2 score of 0.929, showing the value of combining multiple assessments to mitigate individual rater variability.

### Classification performance

The ROC curves demonstrated robust classification performance across all tendon forms, with the general flat tendon achieving the steepest curve, indicating clear separability from other classes (**Supplementary material Figure S5**). A consistently high area under the curve (ROC-AUC = 0.89) confirms that the classification effectively differentiates between tendon forms. It suggests that the classification system provides reliable assessment. The high sensitivity and specificity observed in the ROC analysis highlight the model’s robustness, with a steep slope at a lower false positive rate, indicating minimal misclassification. The cumulative one-versus-rest approach, well suited for these data, ensured that we evaluated classification performance in a clinically meaningful way rather than treating each category as an independent class.

The classification system demonstrated strong overall performance, with an accuracy of 0.83 and a macro-averaged F1 score of 0.83, indicating well-balanced performance across the tendon categories. The macro-averaged precision and recall were 0.84 and 0.83, respectively. The area under the receiver operating characteristic curve was 0.89. A null accuracy of 0.31 confirmed that the classification system substantially outperformed random chance.

### Precision-recall analysis

We used a precision-recall curve to evaluate classification performance; it demonstrates the trade-off between recall and precision. The analysis revealed differences in precision, recall, and the F1 score across the tendon forms. We observed higher F1 scores (**Supplementary material Figure S6**) for tendon forms with more distinct shapes, whereas lower F1 scores reflected greater uncertainty in classification. The high area under the precision-recall curve (0.83) indicates good performance in correctly identifying tendon forms while reducing false positives.

### Error analysis and misclassification trends

We generated a confusion matrix for individual raters (**Supplementary material Figure S7**) to complement Fleiss’ kappa and to provide a more detailed analysis of the misclassifications. The most common error – misclassifying the oval tendon as general flat with a medial bulge (4.76%) – occurred almost twice as much as the other errors (Fig. 3). The confusion matrix shows the frequency with which raters classified tendons as another type, indicating both areas of agreement and misclassification. The diagonal of the matrix, representing the correctly classified tendon forms, was consistently high, suggesting that raters generally agreed with the gold standard (the consensus between two musculoskeletal radiologists).

**Fig. 3.**
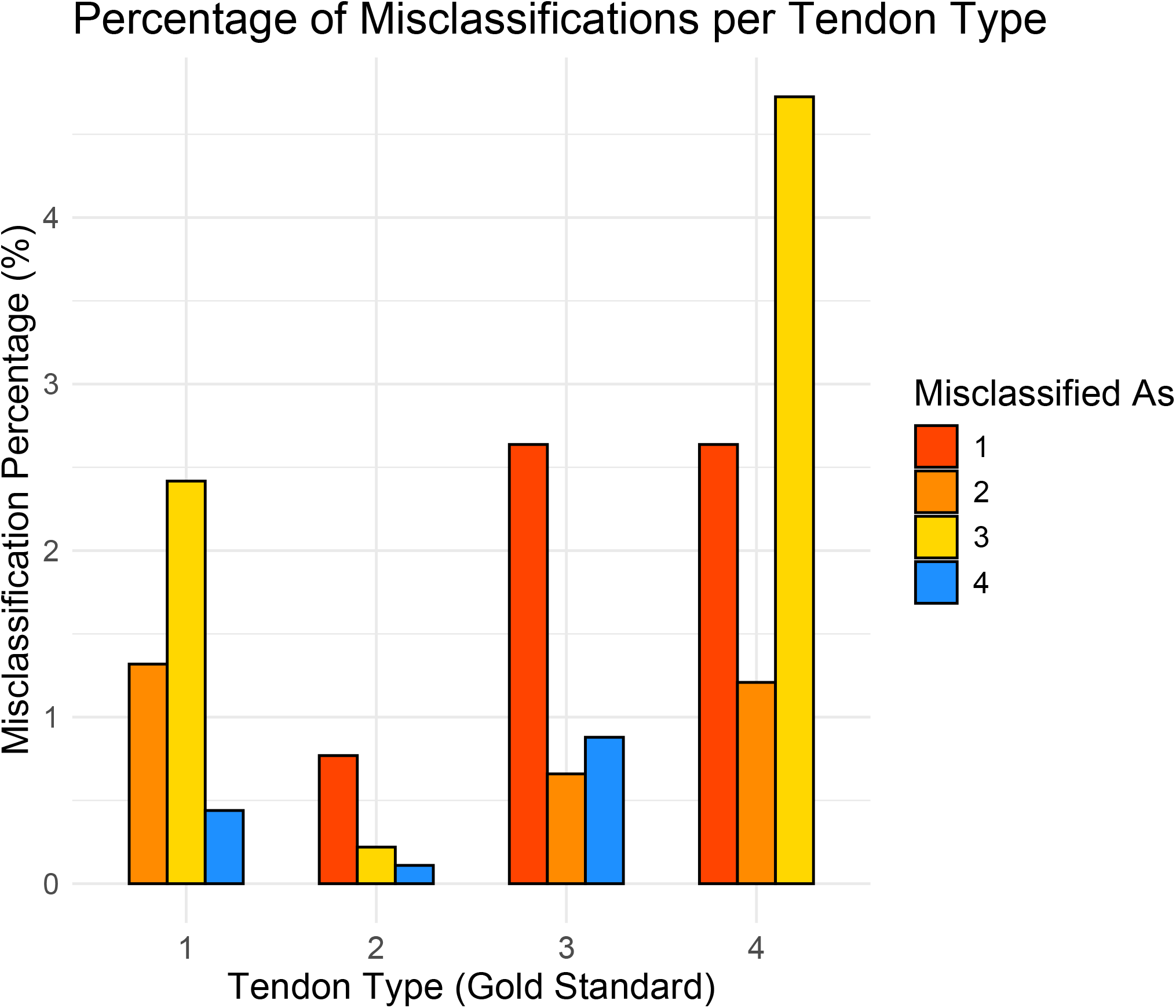
Bar chart showing the percentage (%) of times each tendon form was classified incorrectly by the raters. Tendon forms: 1, general flat; 2, flat with a lateral bulge; 3, flat with a medial bulge; and 4, oval tendon

## DISCUSSION

The most important finding of this study is that the proposed MRI-based classification system for peroneus brevis tendon morphology is reliable and reproducible, with substantial inter-rater agreement. MRI plays a central role in sports imaging, offering detailed assessment of soft tissue structures such as tendons. In athletes with lateral ankle pain or instability, abnormalities in the cross-sectional shape of the peroneus brevis tendon may indicate split tears which are much often than lacerations[14]. This classification standardises the way we assess tendon shape on MRI, helping clinicians distinguish normal variation from early pathological changes. In a sports setting, this can improve diagnostic accuracy by drawing attention to subtle forms linked to split tears, reducing the risk of missed injuries and enabling more informed treatment planning and safer return-to-play decisions. It also offers a reliable framework for future sport imaging research aimed at better understanding the risk factors of split tears.

The substantial agreement based on Fleiss’ kappa suggests that, overall, the raters were able to reliably classify the tendon forms according to the consensus. The consistently higher Gwet’s AC1 values may highlight the limitations of Cohen’s Kappa in cases with uneven category distributions [24]. These findings suggest that individual raters maintained consistency in their assessments, reinforcing the reliability of the classification system. Minor discrepancies may be related to borderline tendon forms or visual assessment, warranting future metric standardisation efforts to further optimise classification consistency. To better understand which types were most frequently confused with one another, we constructed a confusion matrix.

We included raters from different professional backgrounds to reflect the real-world multidisciplinary setting in which athletes with peroneal tendon injuries are assessed. Although our rater group did not include any orthopaedic surgeons, it did include two physiotherapists, offering a balanced mix of raters. Had we included only experienced radiologists, inter-rater agreement might have been higher [25]; however, the findings would have been less generalisable to everyday clinical practice in sport medicine, where patients are managed by multidisciplinary teams and MRI is interpreted by different healthcare professionals.

Some previous studies have referred to the different forms of the peroneus brevis tendon on transverse cross section, but no validated classification exists. Authors have mentioned that a chevron or bisected shape of the peroneus brevis tendon [8] may represent risk factors to split rupture [26]. Overly complex descriptions of tendon shape can lead to misunderstandings.

Therefore, it was necessary to develop a relatively simple classification that still captures the complexity of anatomical variation. To our knowledge, this is the first validated classification of peroneus brevis tendon morphology at the lateral malleolus. We choose this level based on clinical observation and the fact that this is the most common location of a peroneus brevis split rupture [1, 27].

While the overall agreement was substantial [23], we observed variability among the raters, reflecting the challenges of distinguishing subtle anatomical differences. The misclassification patterns seen in the confusion matrix shows the limitations of the visual assessment [28]. The confusion matrix analysis revealed that the oval tendon and the general flat tendon with a medial bulge were frequently misclassified, indicating some similarity in visual classification. This overlap contributed to lower inter-rater agreement and highlights the diagnostic challenges faced in sports medicine when evaluating subtle tendon pathology.

Despite anatomical variations, our analysis demonstrated that the classification remains reliable even when radiologists represent a minority within the group of raters. Multicentre studies with larger and more diverse samples would help confirm whether these misclassification patterns are generalisable across different patient populations. Nevertheless, we determined the sample size prior to commencing the study to ensure we had adequate statistical power and that our results are valid.

Our raters did not perform tendon measurements, as we deliberately chose a purely visual assessment method to reflect real-world clinical scenarios where decisions must be time effective. We hypothesise that, in clinical practice, most raters rely on visual evaluation, occasionally supported by measurements. It is possible that using measurements could have further improved agreement. We think that in the future, the use of artificial intelligence may be used to classify tendon types. However, at present, no such solution exists.

The bootstrapping analysis also highlighted the stability of Fleiss’ kappa, confirming the robustness of our classification system [29]. The narrow confidence intervals and similar Cohen’s kappa and Gwet’s AC1 indicate that the observed level of agreement is unlikely to be due to sampling variability and can be generalised beyond our study cohort. While Cohen’s kappa is widely used in agreement radiological studies, it is known to be sensitive to prevalence imbalances, resulting in the kappa paradox [30]. Gwet’s AC1 is a prevalence- and bias-adjusted kappa statistic [31]. The strong correlation between these two agreement measures confirms the consistency and robustness of the proposed classification system, supporting its potential for additional studies and clinical implementation.

The overlap in the ROC curves suggests some degree of uncertainty in classification between similar tendon forms, which we believe could be further refined with measurements or machine learning models. Future studies should investigate the performance of this classification system across other imaging modalities, including ultrasound, and in diverse international settings.

Despite the strengths of this study, several limitations should be acknowledged. Our classification is based solely on transverse section on magnetic resonance imaging without surgical correlation. Our classification is visual, and we hypothesise that integrating quantitative morphometric analysis could further refine the classification system and improve reproducibility. Our material is based on evaluation of the magnetic resonance examinations performed on clinical indications. Additional prospective studies are warranted for external validation and continued development of the classification system in a more quantitative direction.

## CONCLUSION

Our classification system for the peroneus brevis tendon proved to be a robust and reliable tool, demonstrating substantial inter-rater agreement and excellent overall performance. Misclassifications were rare and mostly limited to tendon types with closely related anatomical features. Its simplicity, practicality, and reproducibility make it well-suited for both clinical and research use. The system’s strength lies in its ability to capture complex anatomical variation in a clear and accessible format. With appropriate rater training, it offers consistent and accurate tendon evaluation, supporting confident diagnosis and informed treatment planning - particularly valuable in athletes, where early detection of tendon tear is essential for guiding return-to-sport decisions.

## DECLARATIONS

## AUTHOR CONTRIBUTIONS

Conceptualization: PS; Methodology: PS, KBD, RZ; Software: PS, RZ; Data Collection: All authors; Data Curation: RZ, PS; Writing – Original Draft: RZ, PS; Writing – Review and Editing: All authors. All authors contributed to previous manuscript versions, and all have read and approved the final manuscript.

## DATA AVAILABILITY STATEMENT

The data supporting this case report are available from the corresponding author on reasonable request, in compliance with the Ethics Committee’s guidelines to ensure patient confidentiality.

## Declarations

Consent to participate The National Ethical Authority approved this study and waived informed consent (Dnr 2024- 07283-02).

## Consent to publish

Not applicable.

