## Supplementary figures and images for "Validation of an MRI-based classification of peroneus brevis tendon morphology: a four-type system with high inter-rater reliability for sports imaging"

### Figure S1

#### Flowchart in the study

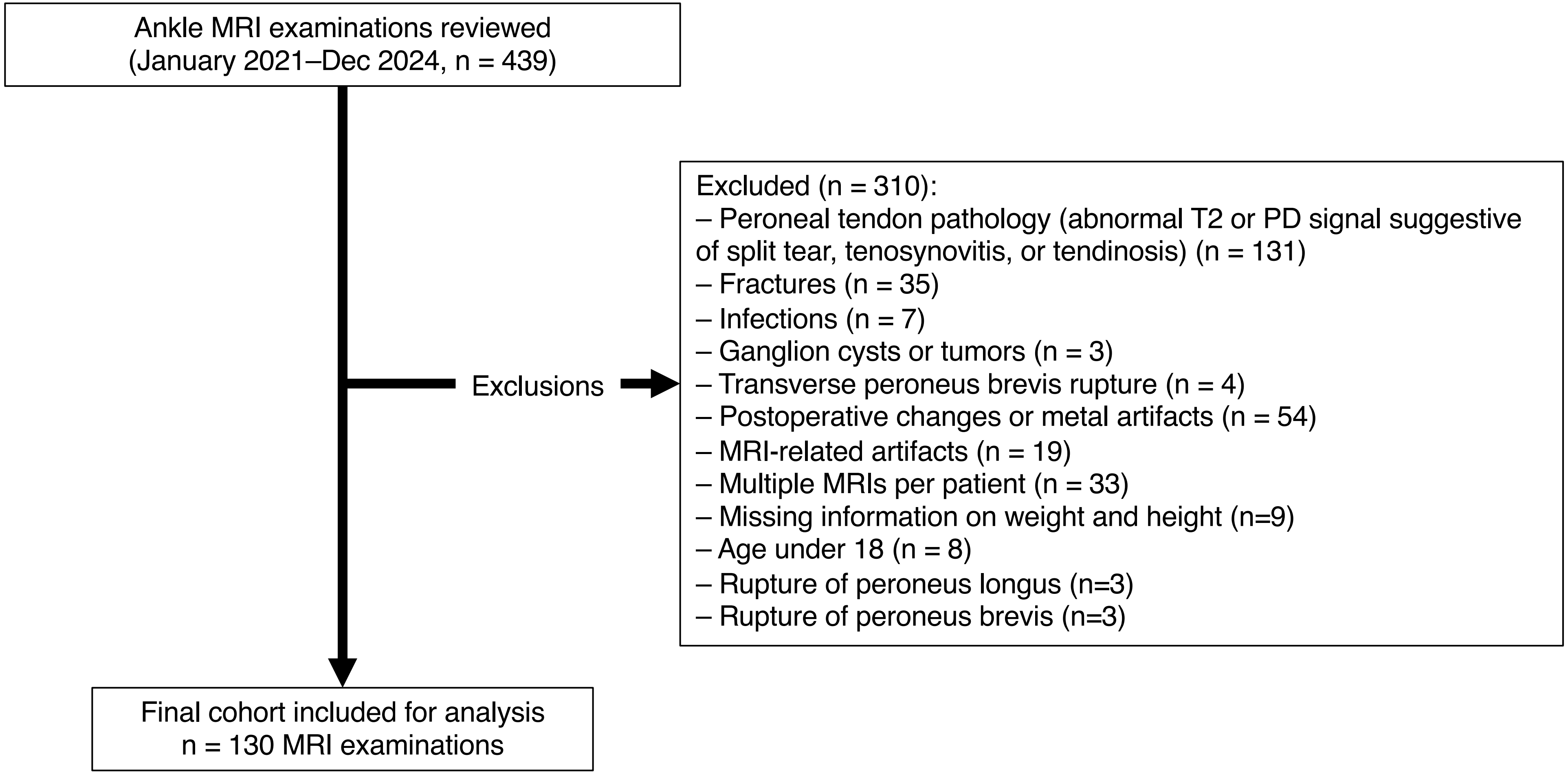

### Figure S2

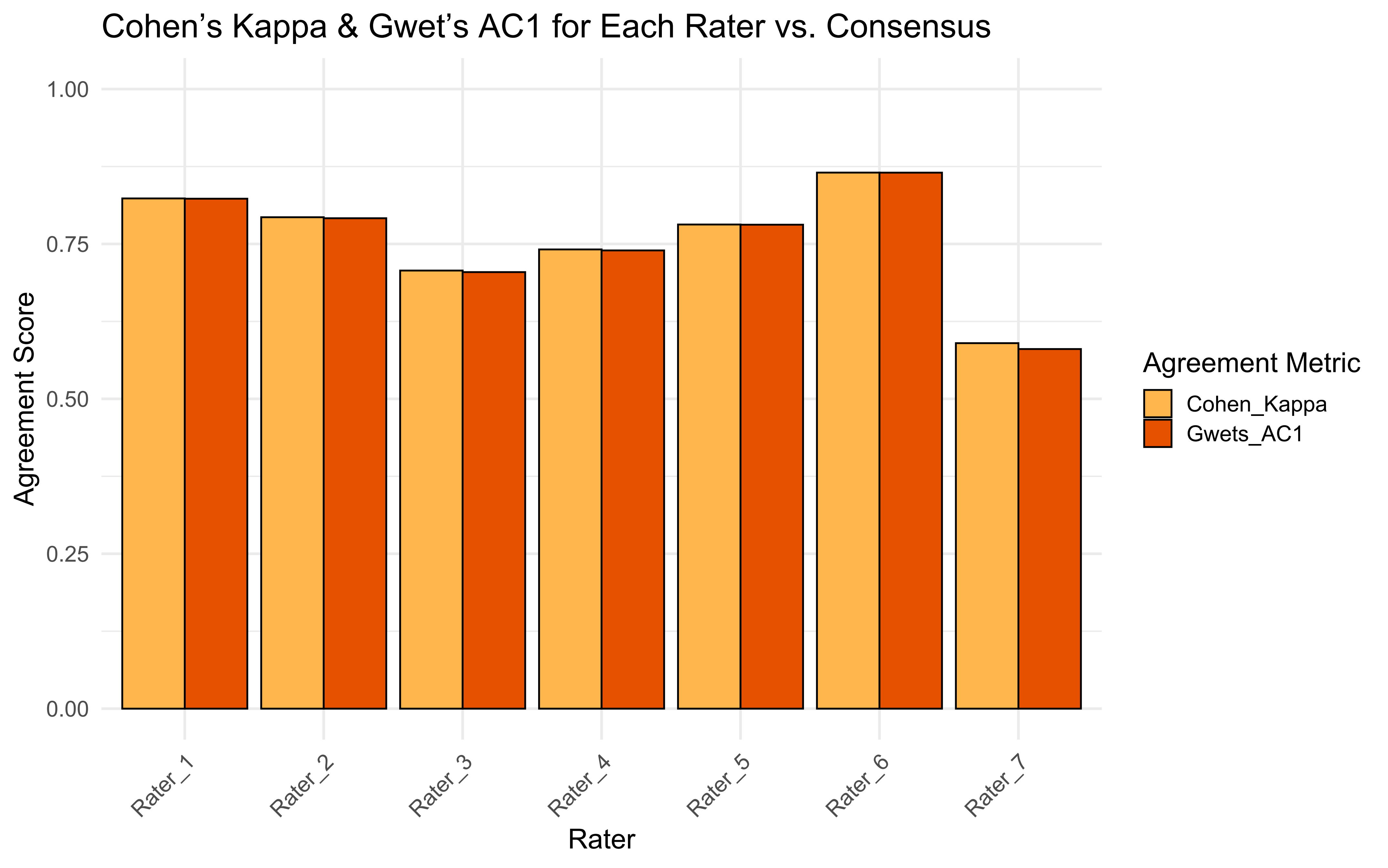

### Figure S3

#### Cohen's Kappa Heatmap Between Raters

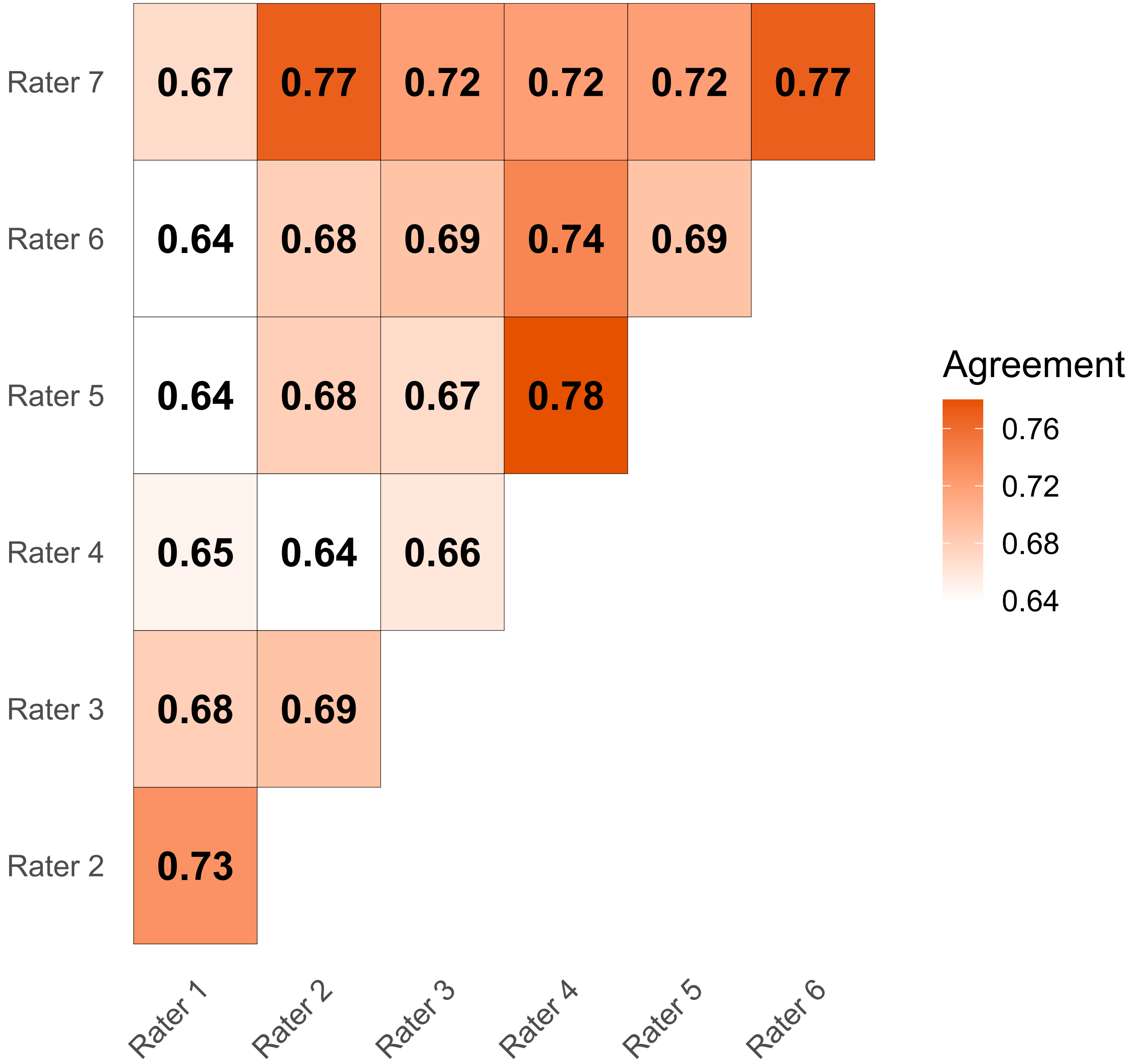

### Figure S4

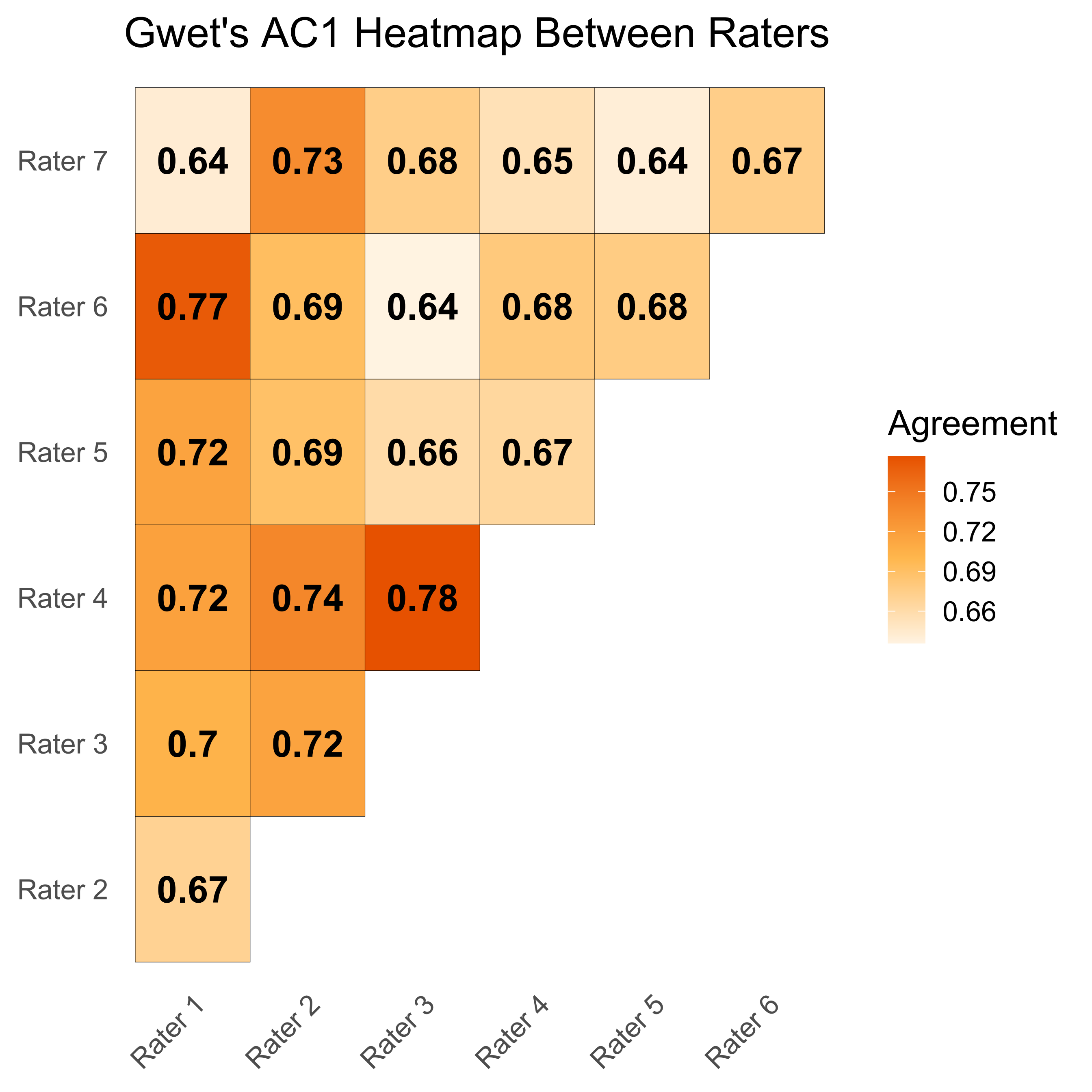

### Figure S5

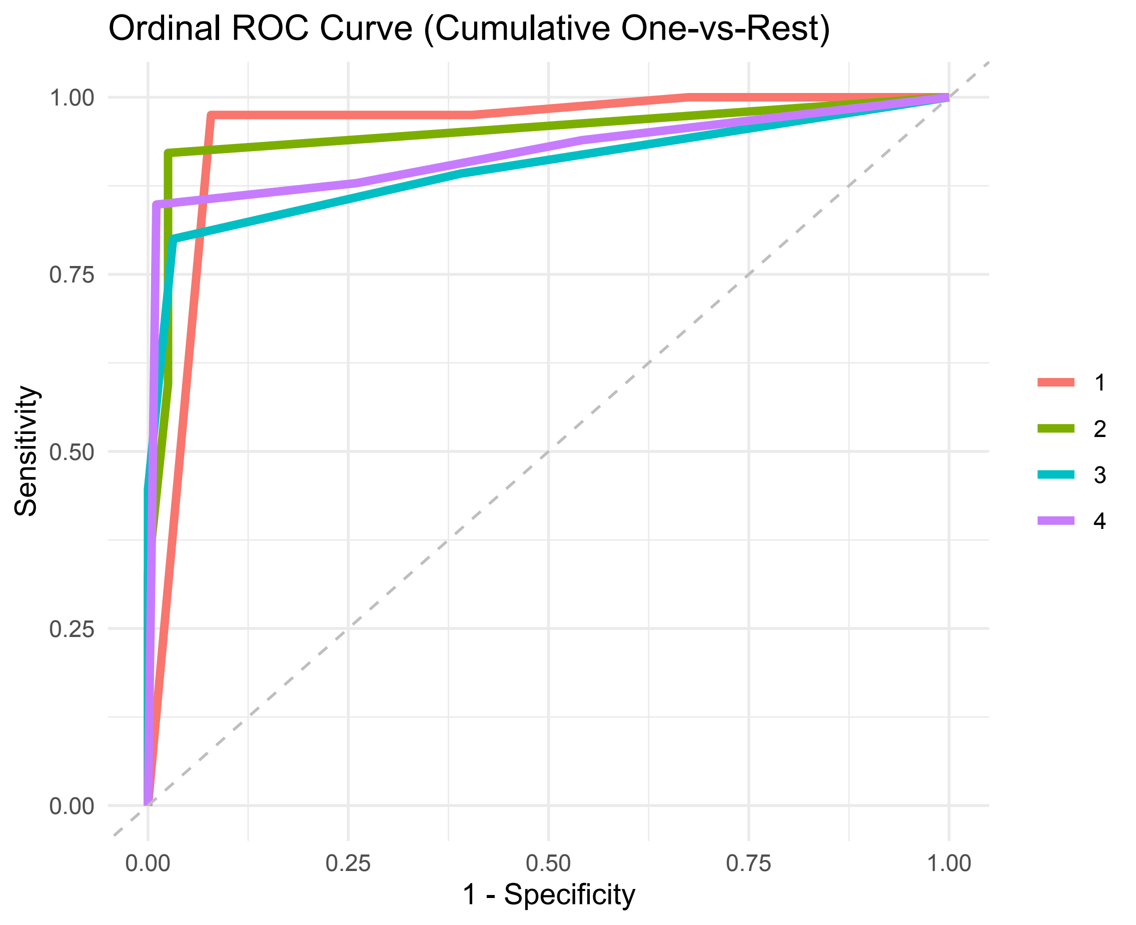

### Figure S6

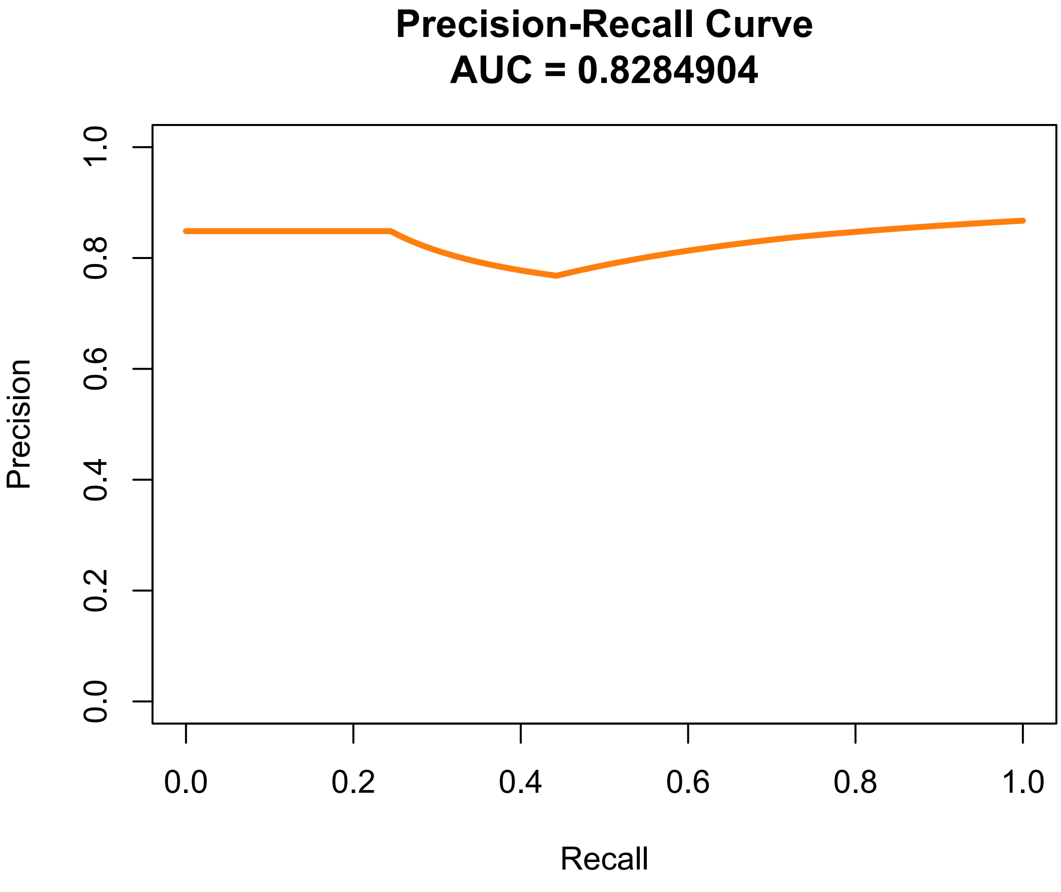

### Figure S7

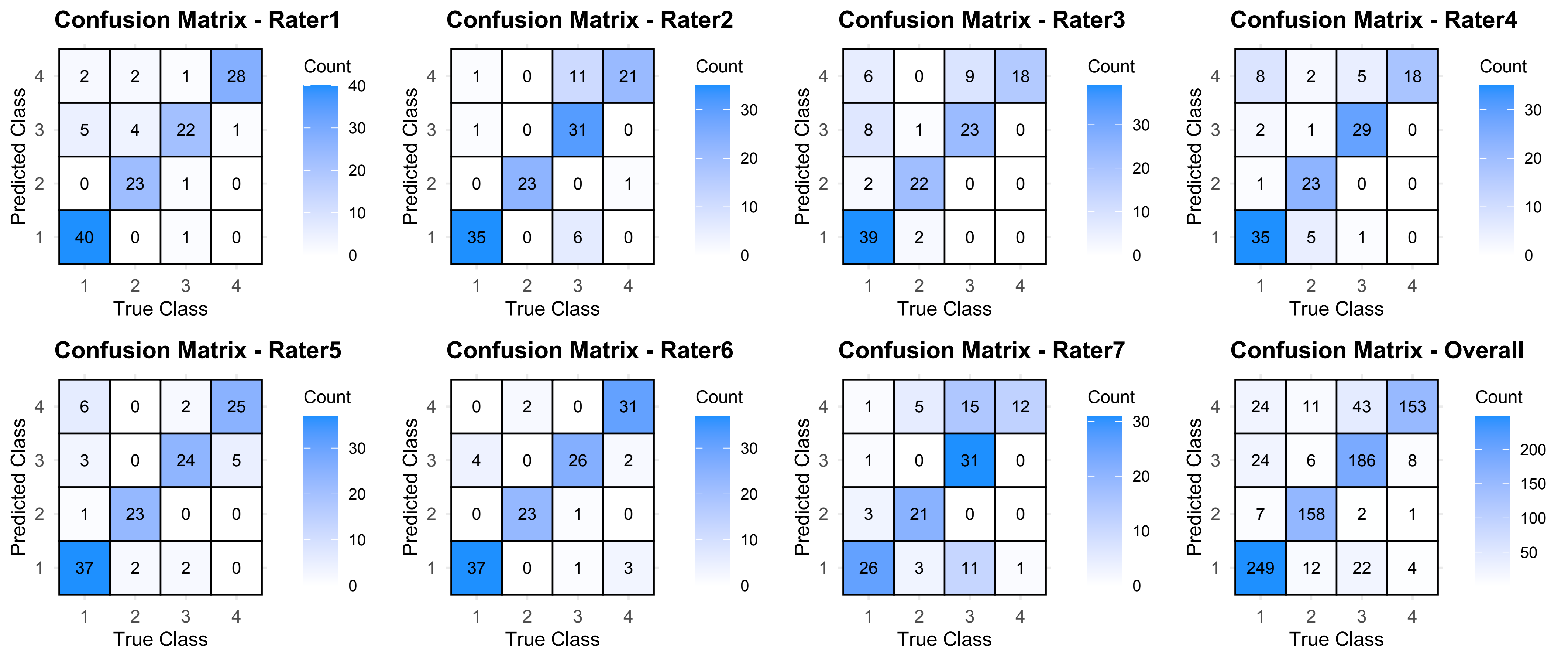
